# Bacterial Sexually Transmitted Infections and Oral HIV Pre-Exposure Prophylaxis Use in HVTN 704/HPTN 085

**DOI:** 10.64898/2026.11.19.26368497

**Authors:** Jorge A. Gallardo-Cartagena, Portia Hunidzarira, Kevin Gillespie, Michal Juraska, Valdiléa G. Veloso, Srilatha Edupuganti, Kelika A. Konda, Alison C. Roxby, Myron S. Cohen, Lawrence Corey, Shelly Karuna, Jorge L. Sanchez

## Abstract

**Objectives:** To describe bacterial STI burden and examine its association with time-varying oral PrEP use among participants in a multinational HIV prevention efficacy trial.

**Methods:** We conducted a post-hoc analysis of HVTN 704/HPTN 085 (ClinicalTrials.gov NCT02716675), a randomized, placebo-controlled study of the VRC01 broadly neutralizing antibody for HIV prevention. Men and transgender persons who have sex with men were enrolled in Brazil, Peru, United States, and Switzerland (April 2016–October 2018). All participants received an enhanced prevention package including STI testing every six months and optional access to oral PrEP. Prevalence and incidence of chlamydia, gonorrhea, and syphilis were described. Cox proportional hazards models estimated associations between time-varying PrEP use and incident STIs.

**Results:** Among 2,687 participants, 27.9% had a bacterial STI at enrollment (chlamydia 11.1%, gonorrhea 10.2%, syphilis 13.1%). Prevalence was higher in Brazil (36.7%) and Peru (34.9%) than in the United States/Switzerland (21.4%). Rectal and oropharyngeal infections predominated. Among participants negative at baseline, incidence of any bacterial STI was 29.4 per 100 person-years (95% CI: 27.2–31.7). Incidence was higher during periods of PrEP use than non-use (HR 1.75, 95% CI: 1.44–2.13), consistent across pathogens.

**Conclusions:** In this multinational HIV prevention trial, bacterial STI burden was high, particularly in Latin America and at extragenital sites. STI incidence was higher during periods of reported oral PrEP use. These findings highlight the importance of integrating multisite STI testing, treatment, counseling, and targeted prevention into HIV prevention services and future efficacy trials.

**Summary:** Participants in HVTN 704/HPTN 085 had high bacterial STI burden, especially in Latin America. Incidence was higher while on PrEP, highlighting need for integrated STI prevention within HIV prevention programs.

## INTRODUCTION

Bacterial sexually transmitted infections (STIs) are a major global public health challenge [1], with disproportionately high incidence among men and transgender persons who have sex with men [2,3]. Over the past two decades, bacterial STIs have increased [1,4] while global HIV incidence has declined with expansion of antiretroviral therapy (i.e., treatment as prevention) and, more recently, pre-exposure prophylaxis (PrEP) [5,6]. The global divergence between rising bacterial STIs and declining HIV incidence has raised questions about how PrEP scale-up, STI testing practices, changing sexual exposure, and broader epidemiologic trends intersect [7,8].

Randomized PrEP trials rarely detected increases in bacterial STIs, possibly because treatment allocation was blinded. Observational studies, however, have reported higher STI diagnoses after PrEP initiation in some settings [9,10], reflecting increased STI testing after entry into PrEP care, changing sexual exposure over time, and PrEP initiation during periods of elevated perceived or actual risk. Most of these studies have relied on pre/post comparisons of STI rates before and after PrEP initiation [11], rather than comparing STI incidence during periods of PrEP use versus non-use. In addition, data from the Americas remain scarce despite recent PrEP scale-up [12,13], particularly analyses linking time-varying PrEP use with systematic screening [14,15].

HVTN 704/HPTN 085 (ClinicalTrials.gov NCT02716675), the Americas/Europe Antibody Mediated Prevention (AMP) study, was a proof-of-concept trial of the broadly neutralizing antibody VRC01 that showed protection against HIV strains sensitive to VRC01 [16]. The study enrolled men and transgender persons with increased likelihood of HIV exposure and provided an enhanced prevention package that included optional oral PrEP and periodic bacterial STI testing [17]. Because oral PrEP was offered but not randomly assigned, uptake varied across participants, allowing examination of the association between time-varying PrEP use and bacterial STI incidence in a large multinational cohort with standardized screening. We used data from HVTN 704/HPTN 085 to describe bacterial STI burden and to estimate the association between time-varying, self-reported oral PrEP use and incident bacterial STIs. These findings may inform STI services in PrEP programs and help plan sexual health support for populations enrolled in future HIV prevention efficacy trials.

## METHODS

### Study Population

HVTN 704/HPTN 085 was a phase 2b randomized, placebo-controlled clinical trial that evaluated VRC01 for HIV prevention in the Americas and Europe among men and transgender persons who have sex with men [16]. The trial enrolled participants at 26 clinical research sites in Brazil, Peru, Switzerland, and the United States between April 2016 and October 2018. For this post-hoc analysis, we included participants in the modified intention-to-treat population, defined as those who received at least one VRC01 or placebo infusion. For each STI outcome, participants were required to have ≥1 scheduled (including enrollment) or interim STI result to contribute to analyses.

### Study Procedures

The parent study protocol was approved by the institutional review boards or ethics committees at each participating site, and all participants provided written informed consent before enrollment. All participants had access to an enhanced HIV prevention package that included condoms, lubricants, HIV prevention counseling, and periodic bacterial STI screening [17]. Testing for bacterial STIs was conducted at scheduled visits every six months through 18 months of follow-up and at interim visits at the discretion of investigators, such as when participants reported symptoms suggestive of an STI.

### PrEP use

Oral PrEP with tenofovir disoproxil fumarate/emtricitabine was available free of charge to all participants, but initiation and continuation were optional and based on participant preference. PrEP use was modeled as a time-varying exposure using self-reported start and stop dates recorded in the concomitant medications log. Person-time between recorded PrEP start and stop dates was classified as “on PrEP”; person-time before initiation, after discontinuation, and among participants who never initiated PrEP was classified as “off PrEP.” In the parent study, dried blood spots were used to assess tenofovir exposure, but those biomarker data were not incorporated into this analysis [18].

### Bacterial STI Testing and Outcomes

Chlamydia testing used nucleic acid amplification tests (NAATs) on urine and rectal samples. Gonorrhea testing used NAATs on urine, rectal, and oropharyngeal samples, although oropharyngeal testing was performed at baseline only. Chlamydia prevalence was defined as a positive NAAT from urine or rectal samples. Gonorrhea prevalence was defined as a positive NAAT from urine, rectal, or oropharyngeal samples. Baseline prevalence of chlamydia and gonorrhea was further described by anatomical site. Syphilis seroprevalence was defined according to local site algorithms using treponemal and non-treponemal assays [19].

The primary outcome was the first incident diagnoses of any bacterial STI. Secondary outcomes were first incident chlamydia, gonorrhea, and syphilis. Participants positive for the outcome at baseline were excluded from the corresponding risk set. Incident chlamydia and gonorrhea were defined as the first NAAT-positive result at any anatomical site during follow-up, and incident syphilis as newly reactive serology during follow-up according to local site algorithms. The trial protocol did not mandate test-of-cure for bacterial STIs, although study sites were responsible for either providing or referring participants for treatment according to local standards. Analyses were restricted to first events because microbiologic clearance was not systematically documented, limiting the distinction between persistent and recurrent infections

### Time at Risk and Censoring

For each STI outcome, person-time accrued from baseline to first incident infection, study completion, dropout, or adjudicated HIV-1 acquisition. Interim STI diagnoses were assigned to the nearest scheduled visit for prevalence summaries. For incidence analyses, events were dated at the first positive specimen collection. The overall annual dropout rate was less than 10% [17].

### Covariates

Sociodemographic and behavioral covariates were assessed at baseline. Age was analyzed as a continuous variable and by categories (≤20, 21–30, and ≥31 years). Race was grouped as Black or African descent, White, or Other, and ethnicity as Hispanic/Latino or not Hispanic/Latino. Gender identity responses included cisgender male, cisgender female, transgender male, transgender female, another self-identified category, or prefer not to answer; for analysis, the latter two categories were combined. Sexual orientation responses included gay/lesbian/homosexual, bisexual, queer, two-spirit, straight/heterosexual, an additional self-described category, not sure, or prefer not to answer. For analysis, sexual orientation was grouped as homosexual, bisexual, heterosexual, or all other responses combined. Geographic region was grouped as United States/Switzerland, Peru, or Brazil. The United States and Switzerland were combined due to the small sample size in Switzerland (n=36), precluding stable country-specific estimates.

Descriptive analyses also considered income, education, and the baseline HIV Behavioral Risk Score (BRS), categorized into tertiles (low, medium, high). The BRS was developed in the parent study using a SuperLearner [20] ensemble as a baseline covariate-predicted probability (on the logit scale) of HIV-1 acquisition diagnosis during primary follow-up. Candidate baseline predictors included clinical site, country, age, race, baseline syphilis, alcohol and drug use, substance use temporally associated with sex, condom use during vaginal or anal sex, and number of vaginal or anal sex partners (Supplementary Table S1). The BRS was constructed using information measured through enrollment and did not include post-enrollment PrEP use or incident STI outcomes.

### Statistical Analysis

Categorical variables were summarized as counts and percentages, and continuous variables as medians with interquartile ranges. Bacterial STI prevalence was estimated at each scheduled study visit and described by sociodemographic characteristics, geographic region, PrEP use status, and BRS tertiles. Baseline prevalence of gonorrhea and chlamydia was further described by anatomical site.

Incidence rates of a first bacterial STI episode were expressed per 100 person-years with exact 95% confidence intervals. Cumulative incidences were estimated using the Kaplan– Meier method. Cox proportional hazards models were used to estimate the association between PrEP use and bacterial STI, with PrEP use modeled as a time-varying covariate as described above. Incident STI events were attributed to the PrEP status of the interval in which they occurred. Models were adjusted for age at enrollment, geographic region, race, ethnicity, and treatment arm (VRC01 vs placebo). The models were intended to estimate the adjusted association between time-varying self-reported PrEP use and STI diagnosis, not the causal effect of PrEP initiation. The HIV BRS was not included because it was developed to predict HIV acquisition rather than bacterial STI incidence and overlapped with several baseline covariates considered in model specification. Post-enrollment behavioral measures were not included because their temporal relationship with PrEP initiation and continuation was not sufficiently resolved to distinguish confounding from mediation. Proportional hazards assumptions were evaluated using score tests of Schoenfeld residuals. Hazard ratios and 95% confidence intervals were calculated using the Wald method; tests were two-sided. Pairwise associations between reporting PrEP use, enrollment age, and baseline BRS were examined using Spearman correlation coefficients to assess potential collinearity before model specification.

## RESULTS

### Study Population

Of 2,701 individuals enrolled in HVTN 704/HPTN 085, 2,687 (99.5%) received at least one study infusion and contributed bacterial STI data to this analysis. Participants were recruited in Brazil (150/2,687, 5.6%), Peru (1,124/2,687, 41.8%), and the United States or Switzerland (1,413/2,687, 52.6%). The majority were aged 21–30 years (1,442/2,687, 53.7%), assigned male sex at birth (2,661/2,687, 99.0%), and self-identified as cisgender men (2,430/2,687, 90.4%) or transgender women (136/2,687, 5.1%). A total of 1,535/2,687 (57.1%) identified as Hispanic or Latino/a, 848/2,687 (31.6%) as White, and 405/2,687 (15.1%) as Black or African American. Baseline demographic and behavioral characteristics have been reported previously [16].

### Baseline Prevalence of Bacterial STIs

At enrollment, 749/2,687 (27.9%) participants tested positive for at least one bacterial STI **(Table 1)**. Chlamydia was diagnosed in 297/2,687 (11.1%) participants, gonorrhea in 273/2,687 (10.2%), and syphilis seropositivity in 353/2,687 (13.1%). Anatomical site-specific testing showed a predominance of extragenital infections. Chlamydia was detected in 240/2,675 (9.0%) participants from rectal samples compared with 80/2,684 (3.0%) from urine samples. For gonorrhea, infections were identified in 154/2,674 (5.8%) participants from rectal samples, 173/2,680 (6.5%) from oropharyngeal samples, and 24/2,684 (0.9%) from urine samples **(Table 2)**.

**Table 1.** Baseline prevalence of bacterial sexually transmitted infections by selected participant characteristics in the HVTN 704/HPTN 085*, 2016-2018 (N=2,687).

| Characteristic | Levels | N | Any bacterial STI<sup>†</sup> | Chlamydia | Gonorrhea | Syphilis seropositivity |
|---|---|---|---|---|---|---|
| Overall |  | 2,687 | 749 (27.9%) | 297 (11.1%) | 273 (10.2%) | 353 (13.1%) |
| Age | ≤20 years | 294 | 105 (35.7%) | 59 (20.1%) | 51 (17.3%) | 40 (13.6%) |
|  | 21–30 years | 1,442 | 429 (29.8%) | 169 (11.7%) | 164 (11.4%) | 187 (13.0%) |
|  | ≥31 years | 951 | 215 (22.6%) | 69 (7.3%) | 58 (6.1%) | 126 (13.2%) |
| Sex assigned at birth | Female | 26 | 2 (7.7%) | 1 (3.8%) | 1 (3.8%) | 0 (0.0%) |
|  | Male | 2,661 | 747 (28.1%) | 296 (11.1%) | 272 (10.2%) | 353 (13.3%) |
| Gender identity | Cisgender male | 2,430 | 665 (27.4%) | 271 (11.2%) | 244 (10.0%) | 306 (12.6%) |
|  | Transgender female | 136 | 48 (35.3%) | 10 (7.4%) | 20 (14.7%) | 28 (20.6%) |
|  | Cisgender female | 43 | 16 (37.2%) | 6 (14.0%) | 5 (11.6%) | 9 (20.9%) |
|  | Transgender male | 16 | 2 (12.5%) | 1 (6.2%) | 1 (6.2%) | 0 (0.0%) |
|  | Another identity/declined<sup>‡</sup> | 62 | 18 (29.0%) | 9 (14.5%) | 3 (4.8%) | 10 (16.1%) |
| Sexual orientation | Homosexual | 2,038 | 588 (28.9%) | 244 (12.0%) | 217 (10.6%) | 273 (13.4%) |
|  | Bisexual | 466 | 120 (25.8%) | 44 (9.4%) | 38 (8.2%) | 61 (13.1%) |
|  | Heterosexual | 40 | 8 (20.0%) | 4 (10.0%) | 3 (7.5%) | 3 (7.5%) |
|  | Other responses<sup>‡</sup> | 143 | 33 (23.1%) | 5 (3.5%) | 15 (10.5%) | 16 (11.2%) |
| Race | Black or African Descent | 405 | 113 (27.9%) | 35 (8.6%) | 32 (7.9%) | 66 (16.3%) |
|  | White | 848 | 176 (20.8%) | 81 (9.6%) | 61 (7.2%) | 65 (7.7%) |
|  | Other | 1,434 | 460 (32.1%) | 181 (12.6%) | 180 (12.6%) | 222 (15.5%) |
| Ethnicity | Hispanic or Latino/a | 1,535 | 498 (32.4%) | 191 (12.4%) | 194 (12.6%) | 249 (16.2%) |
|  | Not Hispanic or Latino/a | 1,152 | 251 (21.8%) | 106 (9.2%) | 79 (6.9%) | 104 (9.0%) |
| Country | US/Switzerland<sup>§</sup> | 1,413 | 302 (21.4%) | 128 (9.1%) | 100 (7.1%) | 125 (8.8%) |
|  | Brazil | 150 | 55 (36.7%) | 11 (7.3%) | 22 (14.7%) | 34 (22.7%) |
|  | Peru | 1,124 | 392 (34.9%) | 158 (14.1%) | 151 (13.4%) | 194 (17.3%) |
| Baseline HIV BRS | High | 896 | 360 (40.2%) | 130 (14.5%) | 142 (15.8%) | 193 (21.5%) |
|  | Medium | 895 | 243 (27.2%) | 96 (10.7%) | 86 (9.6%) | 115 (12.8%) |
|  | Low | 896 | 146 (16.3%) | 71 (7.9%) | 45 (5.0%) | 45 (5.0%) |
| PrEP use at enrollment | Yes | 440 | 92 (20.9%) | 37 (8.4%) | 32 (7.3%) | 40 (9.1%) |
|  | No | 2247 | 657 (29.2%) | 260 (11.6%) | 241 (10.7%) | 313 (13.9%) |
| Treatment arm | VRC01 | 1,789 | 515 (28.8%) | 200 (11.2%) | 199 (11.1%) | 232 (13.0%) |
|  | Placebo | 898 | 234 (26.1%) | 97 (10.8%) | 74 (8.2%) | 121 (13.5%) |

STI = sexually transmitted infection; BRS = HIV Behavioral Risk Score, developed in the parent study to estimate probability of HIV-1 acquisition diagnosis during primary follow-up based on baseline demographic, clinical, and behavioral predictors (Supplementary Table S1), and categorized into tertiles (low, medium, high); PrEP = HIV pre-exposure prophylaxis, assessed by self-report at baseline.
\*Includes all participants who received ≥1 study infusion and had STI results available at enrollment. Prevalence is summarized as n (%).
<sup>†</sup>Any bacterial STI includes chlamydia, gonorrhea, or syphilis. Chlamydia and gonorrhea prevalence were defined by a positive nucleic acid amplification test result from urine, rectal, or oropharyngeal (gonorrhea only) samples at baseline. Syphilis seropositivity was defined according to local algorithms using treponemal and non-treponemal assays.
<sup>‡</sup>Gender identity and sexual orientation were self-reported. “Another identity/declined” combines participants who selected an additional self-described gender identity or chose not to disclose their gender identity. “Other responses” combines sexual orientation responses other than homosexual, bisexual, or heterosexual (including queer, two-spirit, an additional self-described category, not sure, or prefer not to answer). Findings should be interpreted with caution because of small numbers in some categories.
<sup>§</sup>United States and Switzerland are combined due to the small sample size in Switzerland (n=36).

**Table 2.** Baseline chlamydia and gonorrhea positivity by anatomical testing site in the HVTN 704/HPTN 085*, 2016–2018 (N=2,687).

| **STI** | **Testing Site<sup>†</sup>** | **Number Tested** | **Number Positive** | **Prevalence** |
|---|---|---|---|---|
| Chlamydia | Urine | 2,684 | 80 | 3.0% |
| Chlamydia | Rectal | 2,675 | 240 | 9.0% |
| Gonorrhea | Urine | 2,684 | 24 | 0.9% |
| Gonorrhea | Rectal | 2,674 | 154 | 5.8% |
| Gonorrhea | Oropharyngeal | 2,680 | 173 | 6.5% |

STI = sexually transmitted infection.
\*Includes all participants who received ≥1 study infusion and had STI results available at baseline. Variability in denominators reflects incomplete sample collection across anatomical sites.
<sup>†</sup>Urogenital infections were assessed using urine samples. Rectal and oropharyngeal infections were assessed using swabs. All tests were nucleic acid amplification tests.

### Subgroup Differences in Baseline Prevalence

Prevalence of any bacterial STI was highest among participants younger than 20 years (105/294, 35.7%), compared with 429/1,442 (29.8%) among those aged 21–30 years and 215/951 (22.6%) among those 31 years or older **(Table 1).** This prevalence increased with higher baseline HIV BRS: 360/896 (40.2%) in the high tertile compared with 146/896 (16.3%) in the low tertile.

By region, baseline prevalence of any bacterial STI was 55/150 (36.7%) in Brazil and 392/1,124 (34.9%) in Peru, compared with 302/1,413 (21.4%) in the United States/Switzerland. Syphilis seroprevalence was more common in Brazil (34/150, 22.7%) and Peru (194/1,124, 17.3%) than in the United States/Switzerland (125/1,413, 8.8%). Chlamydia prevalence was highest in Peru (158/1,124, 14.1%), and gonorrhea prevalence was similar in Brazil (22/150, 14.7%) and Peru (151/1,124, 13.4%).

Among self-reported gender identity groups, prevalence of any bacterial STI was higher among transgender women (48/136, 35.3%) than among cisgender men (665/2,430, 27.4%). A small number of participants self-identified as cisgender women (n=43) or transgender men (n=16).

### Longitudinal Prevalence Trends

Across scheduled visits, chlamydia and gonorrhea prevalence were numerically lower at later visits, whereas syphilis seroprevalence remained similar or higher, consistent with persistence of treponemal reactivity after infection; these visit-level estimates were descriptive **(Table 3)**.

**Table 3.** Prevalence of bacterial sexually transmitted infections at scheduled study visits in HVTN 704/HPTN 085*, 2016–2018 (N=2,687).

| **Visit (month)** | **N** | **Any bacterial STI<sup>†</sup>** | **Chlamydia** | **Gonorrhea** | **Syphilis seropositivity** |
|---|---|---|---|---|---|
| Baseline | 2,687 | 749 (27.9%) | 297 (11.1%) | 273 (10.2%) | 353 (13.1%) |
| Month 6 | 2,437 | 582 (23.9%) | 202 (8.3%) | 131 (5.4%) | 341 (14.0%) |
| Month 12 | 2,304 | 571 (24.8%) | 185 (8.0%) | 117 (5.1%) | 339 (14.7%) |
| Month 18 | 2,175 | 519 (23.9%) | 154 (7.1%) | 102 (4.7%) | 335 (15.4%) |

STI = sexually transmitted infection.
\*Includes all participants who received ≥1 study infusion and had STI results available at the indicated visit. Prevalence is summarized as n (%).
<sup>†</sup>Any bacterial STI includes chlamydia, gonorrhea, or syphilis. Chlamydia and gonorrhea prevalence were defined by a positive nucleic acid amplification test results from available anatomical sites at each visit; oropharyngeal gonorrhea testing was performed at baseline only. Syphilis seropositivity was defined according to local site algorithms using treponemal and non-treponemal assays. Visit-level estimates are descriptive; syphilis seropositivity may persist after infection and treatment.

### Incidence of Bacterial STIs

Among participants who were negative at baseline, the overall incidence of the first occurrence of any bacterial STI was 29.4 infections per 100 person-years (95% CI: 27.2– 31.7). By pathogen, incidence was highest for chlamydia (16.3 per 100 person-years, 95% CI: 14.7–18.0), followed by gonorrhea (12.8 per 100 person-years, 95% CI: 11.4–14.3), and syphilis (6.9 per 100 person-years, 95% CI: 5.9–7.9).

### PrEP Use and Bacterial STI Outcomes

As previously described, PrEP uptake differed by country (5.3% in Peru, 38.2% in Switzerland, 54.3% in the United States, and 83.2% in Brazil), and participants reporting PrEP use had lower median baseline HIV BRS than non-users (median [range]: PrEP initiators, −0.4 [−2.2 to 3.1]; PrEP non-initiators 0.5 [−2.2 to 3.9]) although the ranges overlapped [18]. Bacterial STI incidence rates were consistently higher while on PrEP compared with off PrEP **(Table 4)**. For any bacterial STI, incidence was 38.5 per 100 person-years (95% CI: 34.3–43.0) during PrEP use versus 24.3 per 100 person-years (95% CI: 21.8–27.0) off PrEP. This pattern was consistent across infections: chlamydia (20.8 on PrEP vs 13.6 off PrEP per 100 person-years), gonorrhea (17.1 vs 10.2), and syphilis (9.1 vs 5.5).

**Table 4.**
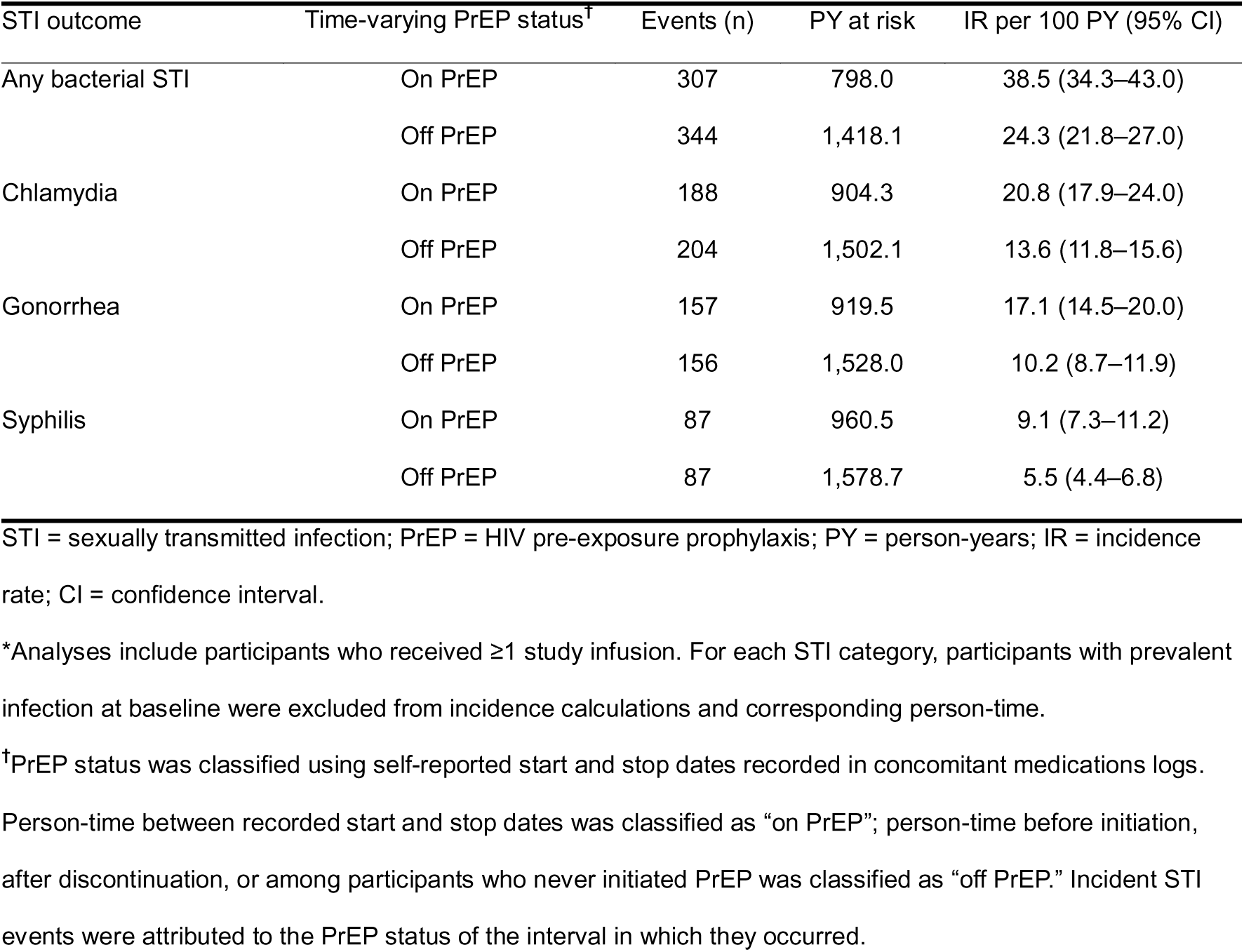
Incidence of first bacterial sexually transmitted infection episode by time-varying oral PrEP use in HVTN 704/HPTN 085*, 2016–2020 (N=2,687).

| STI outcome | Time-varying PrEP status<sup>†</sup> | Events (n) | PY at risk | IR per 100 PY (95% CI) |
|---|---|---|---|---|
| Any bacterial STI | On PrEP | 307 | 798.0 | 38.5 (34.3–43.0) |
|  | Off PrEP | 344 | 1,418.1 | 24.3 (21.8–27.0) |
| Chlamydia | On PrEP | 188 | 904.3 | 20.8 (17.9–24.0) |
|  | Off PrEP | 204 | 1,502.1 | 13.6 (11.8–15.6) |
| Gonorrhea | On PrEP | 157 | 919.5 | 17.1 (14.5–20.0) |
|  | Off PrEP | 156 | 1,528.0 | 10.2 (8.7–11.9) |
| Syphilis | On PrEP | 87 | 960.5 | 9.1 (7.3–11.2) |
|  | Off PrEP | 87 | 1,578.7 | 5.5 (4.4–6.8) |

STI = sexually transmitted infection; PrEP = HIV pre-exposure prophylaxis; PY = person-years; IR = incidence rate; CI = confidence interval.
\*Analyses include participants who received ≥1 study infusion. For each STI category, participants with prevalent infection at baseline were excluded from incidence calculations and corresponding person-time.
<sup>†</sup>PrEP status was classified using self-reported start and stop dates recorded in concomitant medications logs. Person-time between recorded start and stop dates was classified as “on PrEP”; person-time before initiation, after discontinuation, or among participants who never initiated PrEP was classified as “off PrEP.” Incident STI events were attributed to the PrEP status of the interval in which they occurred.

In multivariable Cox models, periods of PrEP use were associated with a higher hazard of being diagnosed with any bacterial STI (HR 1.75, 95% CI: 1.44–2.13). Hazard ratios were similar across first occurrences of pathogen-specific infections: chlamydia (HR 1.69, 95% CI: 1.31–2.17), gonorrhea (HR 1.79, 95% CI: 1.35–2.38), and syphilis (HR 1.91, 95% CI: 1.29–2.84) **(Figure 1).**

**Figure 1.**
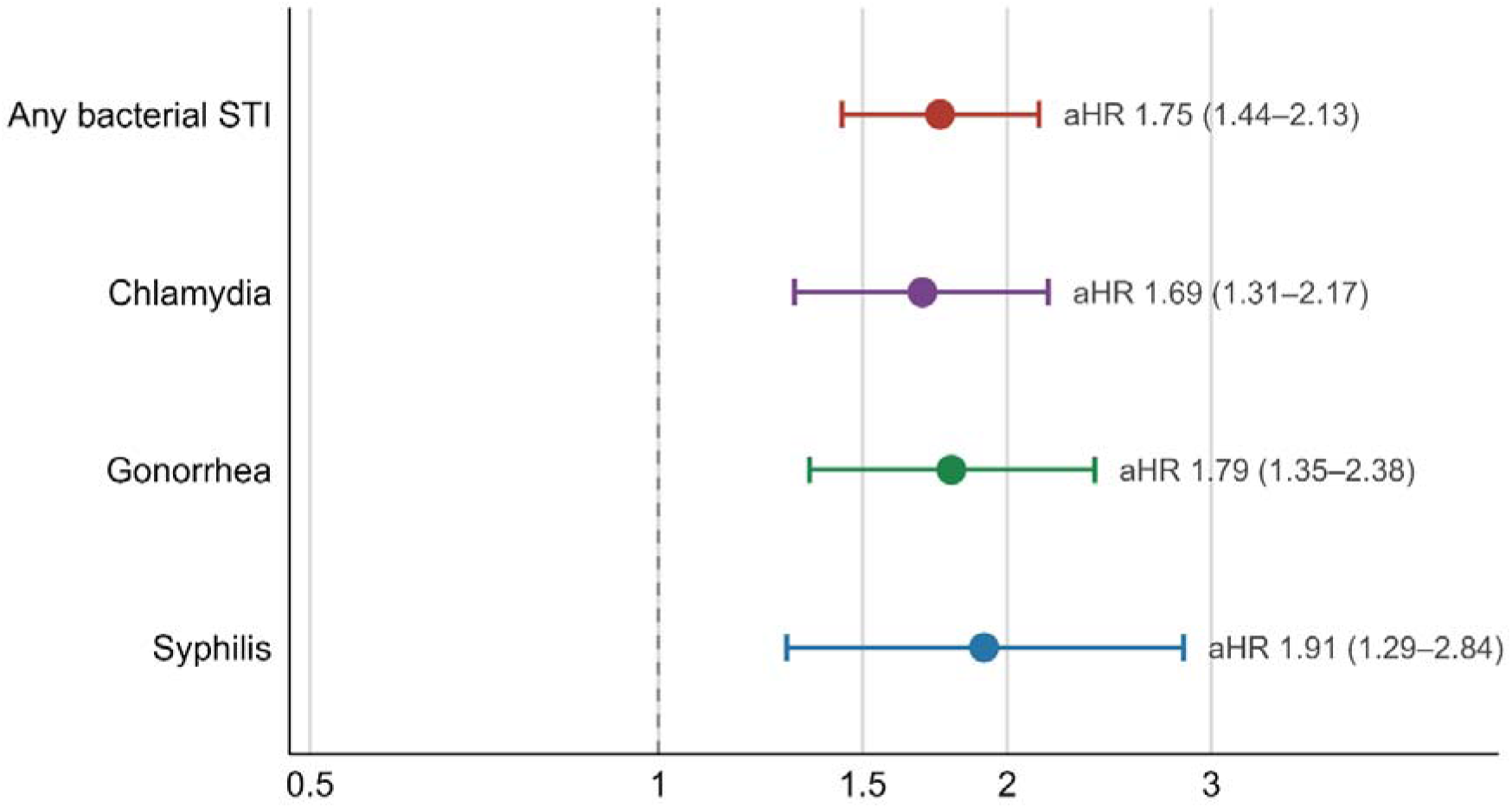
Adjusted hazard ratios (aHRs) and 95% confidence intervals for the association between PrEP use and incident bacterial sexually transmitted infections (STIs) in HVTN 704/HPTN 085, 2016–2020 (N=2,687). Estimates were derived from multivariable Cox proportional-hazards models with time-varying PrEP exposure, adjusted for age at enrollment, geographic region, race, ethnicity, and treatment arm.

## DISCUSSION

In this post-hoc analysis of HVTN 704/HPTN 085, we found a high burden of bacterial STIs among men and transgender persons who have sex with men, with higher incidence observed during periods of PrEP use. At enrollment, over one in four participants had at least one bacterial STI, highlighting the sexual health needs of populations enrolled in HIV prevention efficacy trials. Extragenital infections accounted for a substantial proportion of chlamydia and gonorrhea diagnoses, underscoring the importance of multisite screening beyond urine-based testing [21–24]. Syphilis seroprevalence was highest in Brazil, while chlamydia and gonorrhea were more frequent in Peru and Brazil, respectively, compared with the United States/Switzerland. These regional differences reflect broader global disparities in STI burden potentially linked to structural and health-system factors [1]. During study follow-up, bacterial STI incidence remained high, with the overall rate approaching 30 per 100 person-years despite access to an enhanced prevention package.

We observed that bacterial STI incidence was higher during periods of PrEP use than during non-use, an association that should be interpreted in the context of optional, self-selected PrEP use. Earlier blinded randomized PrEP trials did not show increases in STI incidence, likely because participants were unaware of treatment allocation and, in some cases, because PrEP efficacy had not yet been established [25]. Under these conditions, participants may have been less likely to modify sexual behaviors based on perceived protection. By contrast, subsequent open-label studies have yielded mixed results. A 2018 meta-analysis pooling unblinded PrEP studies (longitudinal observational cohorts, open-label clinical trials, and nonblinded randomized clinical trials) found increased odds of STI diagnosis among PrEP users, particularly in studies conducted after 2016, when PrEP effectiveness had become more widely recognized [9]. Several studies also reported STI increases following PrEP initiation [26–30]. In the Australian PrEPX study, STI incidence rose after initiation compared with the year prior, even after adjusting for testing frequency [29]. Similarly, national surveillance data from Australia showed that syphilis incidence increased steadily during widespread PrEP roll-out [30].

Data from other cohorts indicate that increases in STIs often preceded PrEP initiation, implying that individuals tend to start PrEP during periods of heightened sexual activity [31–33]. For example, in EPIC-NSW [31], STI positivity was stable during PrEP use but had been increasing beforehand, and a Danish before–after study observed a 35% rise in STI incidence beginning 10–20 weeks before PrEP start [32]. Likewise, a long-running observational cohort of MSM in the Netherlands found that PrEP initiators were already on trajectories of increasing number of partners, condomless sex, sexualized drug use, and STIs prior to initiation, which continued after PrEP start compared with matched controls [33]. By contrast, other studies reported no increase or even stabilization despite high background incidence [30,34,35]. In the Amsterdam PrEP demonstration project, STI rates remained persistently high but stable over several years despite reductions in partner numbers and condomless sex [34]. In national surveillance data from Australia, while syphilis incidence continued to rise, chlamydia and gonorrhea declined after early peaks [30], and in Switzerland, incidence of all bacterial STIs actually declined over time [35].

Most recently, a meta-analysis of 23 studies and more than 11,000 participants estimated significant increases in STI occurrence after PrEP initiation, particularly for chlamydia and gonorrhea, with no substantial changes for syphilis [11]. Our findings of higher STI incidence during periods of PrEP use are generally consistent with these results. However, the meta-analysis primarily relied on pre–post comparisons within PrEP cohorts, often with heterogeneous testing schedules and no concurrent control group. In contrast, our analysis leveraged a multinational clinical trial in which all participants underwent standardized STI testing every six months regardless of PrEP use. Because PrEP uptake was limited and varied markedly across countries [18], the cohort included both PrEP users and non-users followed contemporaneously. By modeling PrEP as a time-varying exposure, we estimated hazard ratios comparing STI incidence during PrEP-use and non-use intervals within the same trial follow-up structure. HVTN 704/HPTN 085 thus provides a distinctive setting to evaluate bacterial STI burden across regions and associations between PrEP use and STI incidence. These findings highlight the importance of addressing STI prevention needs alongside HIV prevention, including within efficacy trials.

Notably, participants reporting PrEP use in the parent study had lower HIV BRS at baseline compared with those who did not report PrEP use [18], despite higher STI incidence during PrEP-use intervals. Because the BRS was designed to predict HIV acquisition rather than bacterial STIs, it may not capture changing sexual exposure or networks. This contrast is consistent with several explanations, including PrEP initiation during periods of greater sexual exposure, behavioral changes after initiation, and residual confounding. Thus, baseline HIV BRS may not adequately capture changes in STI vulnerability during follow-up.

Consistent with findings from many PrEP studies, bacterial STI burden remained high in a setting where standard prevention services, including counseling, condoms, and routine screening, were available. Prior work suggests that much of the bacterial STI burden is concentrated within a subset of PrEP users, likely linked through sexual networks with higher background prevalence [14,30,36,37]. In this context, increases in STIs after PrEP initiation may reflect greater sexual exposure, changing partner networks, or changes in perceived HIV risk. However, bacterial STIs have important clinical and immunologic consequences, including mucosal inflammation, even when asymptomatic, and may increase susceptibility to HIV acquisition and transmission [38]. Although timely diagnosis and treatment mitigate these risks, incomplete treatment and recurrent exposure remain challenges. These findings support additional prevention tools, including doxycycline post-exposure prophylaxis [39–44], which has shown high acceptability among potential users and healthcare providers [45,46], as well as future STI vaccines. Although routine screening for asymptomatic gonorrhea and chlamydia remains recommended in several settings, its optimal frequency and balance of benefits and harms, including antimicrobial use, remain debated [47–49]. Targeted prevention strategies may improve efficiency, particularly in resource-limited settings such as Latin America, but should not exclude individuals facing barriers to screening and care.

Our study has several limitations. Although specimens were collected prospectively, optional and self-selected PrEP use left potential residual confounding by time-varying sexual activity, partner number, condom use, sexual networks, and event-driven PrEP use. Higher STI incidence during PrEP-use intervals should therefore be interpreted as an association with self-reported PrEP use, not as evidence of a direct effect of PrEP on bacterial STI acquisition. The HIV BRS was developed to predict HIV acquisition and may not fully capture vulnerability to bacterial STIs. PrEP use was self-reported, and biomarkers of tenofovir exposure collected in the parent study were not incorporated in this analysis, which may have introduced exposure misclassification. Gender identity was self-reported, and estimates for some gender identity groups should be interpreted cautiously because of small sample sizes. Because syphilis testing algorithms varied across sites and titers were not available for this analysis, syphilis incidence was restricted to participants without baseline seropositivity; reinfections among participants with prior syphilis could not be evaluated. We also did not differentiate symptomatic from asymptomatic bacterial STIs, limiting inference about the extent to which routine screening identified otherwise undetected infections. Oropharyngeal testing was limited to gonorrhea and was performed only at baseline. The protocol did not mandate test-of-cure, limiting differentiation of persistent infections from reinfections for chlamydia and gonorrhea. Six-month scheduled testing and discretionary interim visits could miss transient infections or infections treated elsewhere, potentially underestimating incidence. Despite adjustment for treatment arm, trial participation may have influenced risk perception or behavior; this selected 2016–2020 cohort may not represent current PrEP populations.

Despite these limitations, this analysis offers several notable strengths. First, it leverages standardized, prospective STI testing within a large, multinational clinical trial. Second, the analysis goes beyond simple pre/post comparisons by leveraging prospectively collected PrEP exposure data within a common trial protocol. Third, inclusion of sites from both high- and middle-income countries contributes valuable data from the Americas, where PrEP expansion is ongoing, but systematic STI screening data outside high-income settings remain scarce. Finally, the high STI burden observed in AMP, also documented in subsequent HIV vaccine efficacy trials such as Mosaico [50], highlights the sexual health needs of populations enrolled in HIV prevention studies. These findings can inform the planning of context-appropriate STI testing, treatment, and prevention services in future efficacy trials, particularly across settings with differing epidemiologic profiles.

In conclusion, bacterial STI burden was high among participants in this multinational HIV prevention trial, particularly in Brazil and Peru, and STI incidence was higher during reported PrEP use than non-use. Our findings support integrating context-appropriate multisite STI testing, treatment, and targeted prevention, including doxycycline post-exposure prophylaxis where appropriate, into HIV prevention programs and future efficacy trials.

## Data availability statement

Deidentified data supporting the primary HVTN 704/HPTN 085 trial publication are publicly available through Harvard Dataverse (https://doi.org/10.7910/DVN/GDESJI).

## Ethics approval statement

The HVTN 704/HPTN 085 study protocol was approved by the institutional review boards or ethics committees at each participating clinical research site. The study was conducted in accordance with applicable ethical and regulatory requirements. The present study is a secondary analysis of data collected under the approved parent protocol.

## Patient consent statement

All participants provided written informed consent before enrollment in HVTN 704/HPTN 085. The present analysis used deidentified study data, and results are reported in aggregate without identifiable participant information.

## Clinical trial registration

The parent HVTN 704/HPTN 085 (Americas/Europe AMP) trial was registered at ClinicalTrials.gov (NCT02716675). The present manuscript reports a post hoc secondary analysis of trial data.

## Acknowledgments

We thank the participants of HVTN 704/HPTN 085, the Protocol Team, and the staff of the 26 participating clinical research sites, including their community engagement and recruitment teams, whose contributions made this study possible. We also thank the Community Advisory Boards at each site for their partnership and support. J.A.G.-C. acknowledges the HVTN Scientific Leadership Development Program for supporting his leadership of this analysis. During manuscript preparation, the authors used ChatGPT (OpenAI) for language editing and refinement. All content was reviewed and approved by the authors, who take full responsibility for the final manuscript.

## Author contributions

Conceptualization: J.A.G.-C., P.H., A.C.R. Methodology: J.A.G.-C., K.G., M.J. Formal analysis: K.G., M.J. Data curation: K.G., M.J. Writing—original draft: J.A.G.-C. Writing—review and editing: all authors. Supervision: A.C.R., K.A.K., J.L.S. Project administration and scientific oversight of the parent HVTN 704/HPTN 085 trial: M.S.C., L.C., S.E., S.K.

