## Supplemental Tables for "Bacterial Sexually Transmitted Infections and Oral HIV Pre-Exposure Prophylaxis Use in HVTN 704/HPTN 085"

| **Supplementary Table S1. SuperLearner composition and baseline predictors contributing to the baseline risk score for HIV-1 acquisition** | | | |
| --- | --- | --- | --- |
| **Learner** | **Screening method** | **SuperLearner weight** | **Predictors contributing to learner*** |
| **SL.mean** | All | **0.428** | None; mean (intercept-only) learner |
| **SL.glm** | Univariate logistic *P*<0.10 | **0.283** | Study site; country; age; race; baseline syphilis; alcohol use frequency; frequency of ≥6 drinks at once; number of alcoholic drinks on a typical drinking day; heroin/prescription painkiller use; history of IV infusion |
| **SL.xgboost** | All | **0.188** | Age; country; baseline syphilis; frequency of ≥6 drinks at once; alcohol use frequency; race; history of IV infusion; age category; study site |
| **SL.glmnet** | All | **0.101** | None; predictor coefficients were shrunk to zero |
| BRS, baseline risk score; GLM, generalized linear model; GLMnet, penalized generalized linear model; XGBoost, extreme gradient boosting.  *Predictors are reported at the variable level rather than individual factor levels. The SuperLearner was fitted using placebo-arm data, and the BRS was derived from the weighted ensemble predictions. Only learner-screen combinations assigned a non-zero SuperLearner weight are shown; the weights sum to 1.00. The univariate logistic screen retained variables with a two-sided Wald-test P<0.10. Additional candidate baseline predictors considered but not retained in a predictor-contributing learner were amphetamine/methamphetamine use; sex within 2 hours of amphetamine/methamphetamine use; cigarette/e-cigarette use; crack cocaine use; sex within 2 hours of crack cocaine use; sex within 2 hours of heroin/prescription painkiller use; injection drug use; marijuana/dagga/K2/spice use; MDMA/Ecstasy use; sex within 2 hours of MDMA/Ecstasy use; sex within 2 hours of marijuana use; other drug use; popper use; sex within 2 hours of popper use; powder cocaine use; sex within 2 hours of powder cocaine use; condom use during vaginal or anal sex; vaginal or anal sex while drunk; and number of vaginal or anal sex partners. Variables with >5% missing values and sparse indicator variables were excluded according to prespecified screening rules; remaining missing values were imputed. | | | |
